# A 16-Month Longitudinal Investigation of Risk and Protective Factors for Mental Health Outcomes Throughout Three National Lockdowns and a Mass Vaccination Campaign: Evidence from a Weighted Israeli Sample During COVID-19

**DOI:** 10.1101/2022.03.18.22272624

**Authors:** Nimrod Hertz-Palmor, Shachar Ruppin, Noam Matalon, Mariela Mosheva, Shirel Dorman-Ilan, Asia Avinir, Ehud Mekori-Domachevsky, Ilanit Hasson-Ohayon, Raz Gross, Doron Gothelf, Itai M. Pessach

## Abstract

**Background:** The COVID-19 pandemic is an ongoing global crisis, with a multitude of factors that affect mental health worldwide. Here, we explore potential predictors for the emergence and maintenance of depression, anxiety, and posttraumatic stress symptoms (PTSS) in the general population in Israel.

**Methods:** Across the span of 16 months, 2,478 people completed a repeated self-report survey which inquired psychiatric symptoms and pandemic related stress factors (PRSF). PRSF were divided into four clusters of environmental stressors: financial, health-related, fatigue and sense of protection by authorities. We applied mixed-effects linear models to assess how each stressor contributes to depression, anxiety and PTSS at each time point, alongside a longitudinal exploration among participants who completed at least two consecutive surveys (*n*=400).

**Results:** Fatigue was the strongest predictor for depression, anxiety and PTSS at all time points (*standardized β* between 0.28-0.60, *p*<.0001), and predicted deterioration overtime (*β* between 0.22-0.36, *p*<.0001). Financial concerns associated with depression and anxiety at all time points (*β* between 0.13- 0.26, *p*<.01), and with their deterioration overtime (*β* between 0.16-0.18, *p*<.0001), while health related concerns were uniquely associated with anxiety and PTSS at all time points (*β* between 0.14-0.29, *p*<.01) and their deterioration (*β* between 0.11-0.16, *p*<.001), but not with depression. Improvement in sense of protection overtime associated with decrease in depression and anxiety (*β* between −0.09 to −0.16, *p*<.01).

**Conclusions:** Our findings accentuate the multitude of risk factors for psychiatric morbidity during COVID-19, and the dynamics in their association with different aspects of psychopathology at various time points.

## 1. Introduction

The COVID-19 pandemic is still an ongoing global crisis, which have so far taken the lives of over 4.9 million people, and have left many others with long-term health conditions (Taquet et al., 2021; World Health Organiztion, 2021). Even among those who were not infected with the virus, many people’s health has still taken a hit due to the indirect effect of COVID-19 on the economy (Brodeur, Gray, Islam, & Bhuiyan, 2021; Ibn-Mohammed et al., 2021), daily routine (Douglas, Katikireddi, Taulbut, McKee, & McCartney, 2020) and mental health worldwide (Pierce, Hope, et al., 2020; Vindegaard & Benros, 2020; Xiong et al., 2020). During its first months, the pandemic had adverse psychological ramifications (Tsamakis et al., 2021), most notably increased rates of anxiety and depression (Aknin et al., 2021; Arad, Shamai-Leshem, & Bar-Haim, 2021; Ebrahimi, Hoffart, & Johnson, 2021; Salari et al., 2020; Tsamakis et al., 2021; Vindegaard & Benros, 2020). However, in most reports this incline in psychopathology returned to normal pre-pandemic rates by mid-2020 (see Varga et al., 2021).

While investigating the emotional impact of the pandemic, concentrating on trajectories of psychiatric disorders at the population level can mask significant heterogeneity in the ways that individuals have been affected, and how different emotional, economic, health-related and social stressors contribute to this heterogeneity. For instance, Aknin et al.(2021) found that many stressors that were associated with depression and anxiety during COVID-19 resembled well-established risk factors for psychiatric morbidity from pre-pandemic times (Browning et al., 2021; Pierce, Hope, et al., 2020; Wang et al., 2021). These include being a female, unemployed or inactive, a member of a minority or marginalized racial group, living in urban areas, belonging to the lowest income quintile, living without a partner, or having a pre-existing medical condition (Aknin et al., 2021). Alongside, Pierce et al. (2020) pinpointed some novel risk factors, that were associated with the rise of psychological distress specifically during COVID-19. These factors include being young (age 18-34) and having young children at home, echoing previous studies on global disasters that emphasize younger age as a risk factor (e.g., Kuwabara et al., 2008). Evidence from Israel, where the present study was conducted, resonates with these conclusions (Horesh, Kapel Lev□Ari, & Hasson□Ohayon, 2020; Lipskaya-Velikovsky, 2021; Oryan, Avinir, Levy, Kodesh, & Elkana, 2021). However, previous studies from Israel were based on small samples, and were neither representative nor weighted, and therefore prone to numerous biases that were widely discussed in the context of mental health research during the pandemic (Pierce, McManus, et al., 2020). Moreover, studies from Israel to date are mostly cross-sectional, and as such they do not provide information on the dynamics of risk and resilience factors overtime, as the circumstances changed dramatically through lockdown orders and mass vaccination.

One of the most widely discussed psychological effects of the pandemic is psychological and physical fatigue, which in the current context is often addressed to as ‘pandemic fatigue’ (Michie, West, & Harvey, 2020). Fatigue is a wide construct that includes feelings of physical or mental exhaustion and depletion of mental resources, which can be attributed to lack of sleep, health conditions (Phillips, 2015) or chronic stress (Weeks, McAuliffe, DuRussel, & Pasquina, 2010). While ‘pandemic fatigue’ was considered a controversial phenomenon in the early days of the pandemic (Reicher & Drury, 2021), especially since many people viewed it as a common excuse for disobeying governmental restrictions, there have been accumulating evidence to its existence ever since, two years into the pandemic (Morgul et al., 2021; Petherick et al., 2021). We have previously demonstrated the substantial contribution of mental exhaustion and sleep difficulties to adverse mental outcomes among healthcare workers (Mosheva et al., 2021, 2020) and relatives of COVID-19 infected patients (Hertz-Palmor et al., 2022) during the early phase of the pandemic in Israel. In both cases, mental exhaustion was the strongest predictor for anxiety, depression, and posttraumatic stress symptoms (PTSS).

In the current study, we expand our previous investigation on the association among depressive symptoms and financial hardships during the first month of the pandemic (Hertz-Palmor et al., 2021), and report findings from the general population in Israel during a 16-month period, stretching from the early days of the pandemic throughout three national lockdowns and a nationwide vaccination campaign. The overarching aims of the current research were to: a) illustrate trends in depression, anxiety, and PTSS during the first 16 months of the COVID-19 pandemic in Israel; b) to explore risk and protective factors for depression, anxiety, and PTSS, and assess their longitudinal dynamics. While these predictive factors can be personal and/or contextual, we focus on four clusters of environmental stressors that are examined as possible predictors, using a designated Pandemic-Related Stress Factors (PRSF) inventory (Imai et al., 2010; Mosheva et al., 2020). To reduce dimensionality in these environmental stressors, we conducted an exploratory factor analysis that converged with the theoretical constructs of the PRSF inventory and concluded in four latent factors: financial, health-related, fatigue, and sense of protection by the authorities. In line with our findings among healthcare workers (Mosheva et al., 2020) and caregivers of COVID-19 patients (Hertz-Palmor et al., 2022), we hypothesized that fatigue would be the strongest predictor for depression, anxiety and PTSS. In line with our previous findings among the general population (Hertz-Palmor et al., 2021), we expected that financial concerns would most strongly associate with depression, and would exceed the association of health-related concerns with depression.

## 2. Methods

### 2.1. Participants, setting and procedure

Participants of this study were Israeli adults (18 years or older), who were invited to participate via social media and instant messaging. On March 17^th^, 2020, the Israeli government has announced a national lockdown. A day later, we launched our first survey (T1) and distributed it via Facebook groups and chain messages on WhatsApp. After completing the survey, the participants were offered to leave their contact information if they wished to be re-contacted for future surveys. The next three surveys were delivered at the end of the first lockdown (T2, April-May 2020) and at the beginning of the second (T3, September-October 2020) and third (T4, December 2020) lockdowns. The fourth time point (T4) was adjacent to the beginning of the national vaccination campaign. The fifth and final survey was conducted at June 2021, 6 months after the beginning of the vaccination campaign (Figure 1). At that time of final survey, there were initial reports regarding the emergence of the delta variant (Shitrit, Zuckerman, Mor, Gottesman, & Chowers, 2021), and obligatory wear of face mask as well as reinstallation of the “Green Pass” restrictions ordinance (Wilf-Miron, Myers, & Saban, 2021) were restored not long after. We conducted two rounds of participants recruitment-cohort 1 (*n=*1,262) was recruited during T1 and was followed throughout T2-T5. Cohort 2 (*n*=1,216) was recruited during T3 and followed throughout T4-T5.

**Figure 1.**
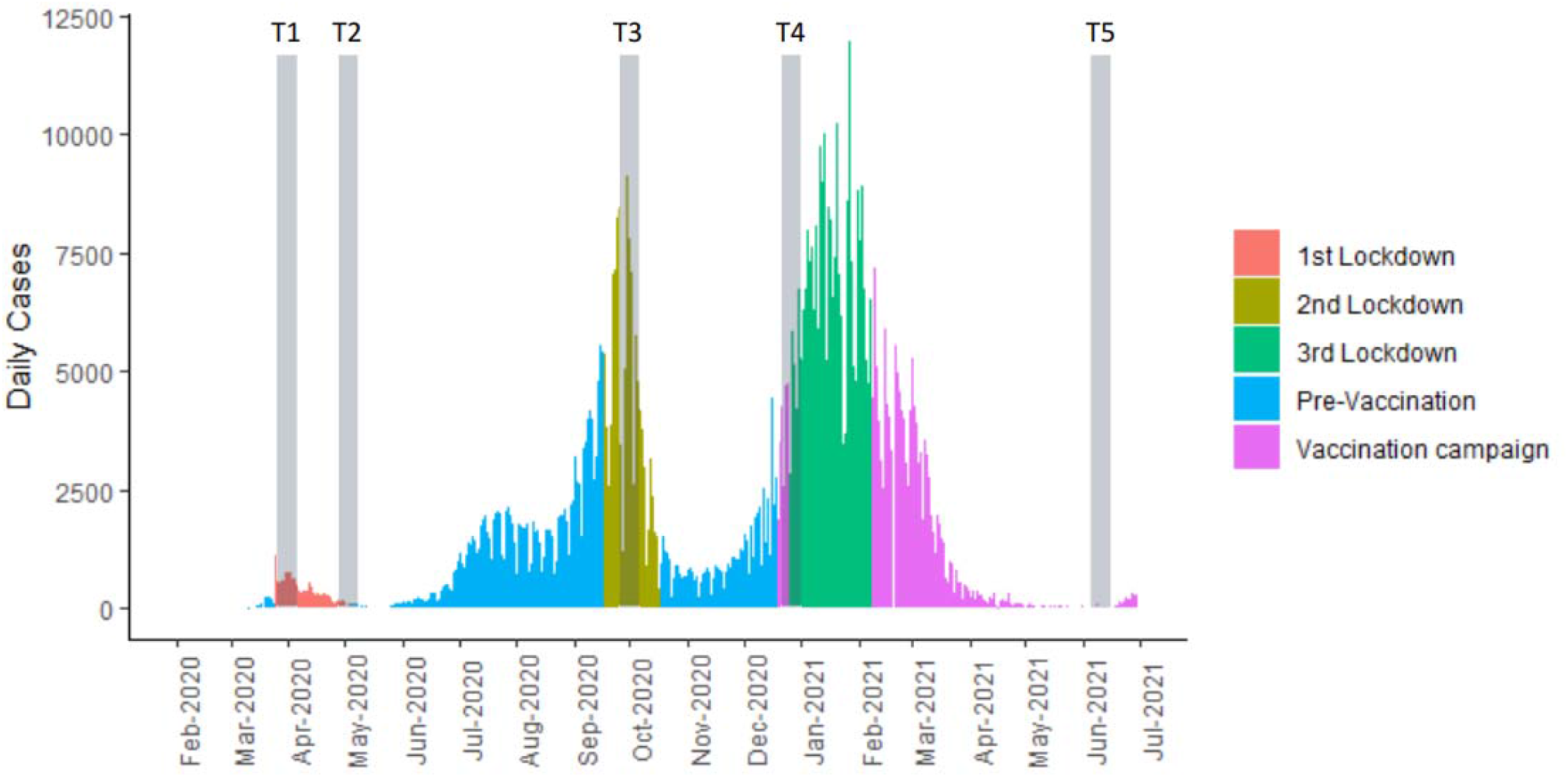
Study timeline

### 2.2. Measures

To measure Depression and Anxiety, we used the well-validated Patient-Reported Outcome Measurement Information System (PROMIS; see www.nihpromis.org), developed by the NIH. PROMIS has good convergent validity with common self-report measurement tools for psychopathology, such as the Patient-Health Questionnare-9 and General-Anxiety-Disorder-7 (Choi, Schalet, Cook, & Cella, 2014; Schalet, Cook, Choi, & Cella, 2014; Schalet et al., 2016; Sunderland, Batterham, Calear, & Carragher, 2018), and it was successfully utilized in previous studies which inquired on mental health in Israel during COVID-19 (Dorman-Ilan et al., 2020; Hertz-Palmor et al., 2021; Matalon et al., 2021; Mosheva et al., 2021, 2020). PROMIS have validated cutoffs for probable depression (Choi et al., 2014) and anxiety (Schalet et al., 2014), equivalent to the common ≥10 cutoff of PHQ-9 and GAD-7. We previously used PROMIS cutoffs to portray the prevalence of depression and anxiety among relatives of COVID-19 patients (Hertz-Palmor et al., 2022). Posttraumatic stress symptoms were screened with the validated Hebrew version of the Primary Care PTSD Screen for DSM-5 (PC-PTSD-5; Spoont et al., 2015). PC-PTSD-5 has a cutoff of ≥3 for probable PTSD. COVID-19 related stressors were assessed with a designated inventory of pandemic-relates stress factors (PRSF), originated by Imai et al. (2010) during the N1H1 pandemic and modified by Mosheva & Hertz-Palmor et al. (2020) to the context of the COVID-19 pandemic. The PRSF inventory includes queries on several relevant stressors on a 4-point Likert scale, and a cutoff of ≥3 for high stress (Mosheva et al., 2020). We inquired after income with a 5-point Likert rating scale, addressing the person’s income with regards to the average wage in Israel (which was extracted from Israel Bureau of Statistics). Actual income loss during the pandemic was assessed on a 5-point Likert scale ranging from “no income loss” to “extreme income loss”. Sociodemographic items were presented at the beginning of the survey. To incentivize recurring participation and survey completion, the participants were presented at the end of each survey with previous findings from older surveys.

### 2.3. Statistical analysis

We use descriptive statistics to present the sample’s characteristics, with respect to the two cohorts that were recruited during the first (cohort 1, March 2020) and second (cohort 2, September 2020) national lockdowns. The temporal dynamics in depression, anxiety, PTSS and PRSF were tested with chi-square tests, using PROMIS, PC-PTSD-5 and PRSF cutoffs.

#### Cross-sectional investigation

To explore risk and protective factors, we conducted three separate mixed-effects linear regressions with depression, anxiety and PTSS scores as the dependent variable in each model. Independent variables included PRSF and the following sociodemographic factors: age, sex, living with a spouse, living with children and income. Participants who completed two or more time points were included as random effects in the models. The time of completing the survey (T1-T5) and the participant’s cohort (1 or 2) were also controlled and modeled as random effects. To avoid overfitting and multicollinearity that may arise from multiple PRSF and their intercorrelations, we used exploratory factor analysis (EFA) with Promax rotation to reduce PRSF dimensionality, and aggregated the scores of clustered items to represent latent factors, which included the following: I) Financial (consisted of financial concerns and income loss), II) Health-related (consisted of anxiety getting infected, anxiety infecting family with COVID-19, lack of knowledge about infeciousity and virulence and lack of knowledge about prevention and protection from the virus), III) Fatigue (consisted of mental exhaustion, physical exhaustion, sleeping difficulties and feelings of social disconnection and being shunned by others), and IV) Sense of protection by the authorities (consisted of feeling protected by the government and local authorities, and feeling protected by the healthcare system). A full description of the items which comprise each cluster and their factor loadings are available at the supplementary materials. Since our interest lay with the effects of PRSF on each time point specifically, we did not include a main effect of PRSF in the model, and instead introduced the PRSF*time point interactions as fixed effects.

#### Longitudinal investigation

To explore the synchronization among changes in stressors and mental health outcomes (i.e., their covariance overtime), we used longitudinal data (i.e., participants who completed at least two consecutive time-points, *n*=400, *observations*=960) to calculate delta measures for depression, anxiety, PTSS and PRSF, by subtracting each measure score at time T from the preceding score at time T-1. For example, the delta measure for depression was calculated as follows:

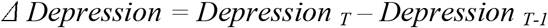

Positive scores in the delta measures represented a rise in depression, anxiety, PTSS or PRSF, while negative scores represented a reduction of symptoms. We repeated our main analysis with the delta scores to assess whether exacerbation (i.e., increase) in mental health outcomes was associated with increased pandemic-related stressors.

To address data biases, we weighted our data to resemble the age and sex distribution in Israel 2020, with data curated from the Israel Central Bureau of Statistics official website (available at https://www.cbs.gov.il/EN/pages/default.aspx). A detailed description of the weighting procedure is available in the supplementary materials of the online version.

In all of our analyses we used a stringent α of .01. Analysis was conducted using the *stats* and *lmerTest* packages in R (Kuznetsova, Brockhoff, & Christensen, 2017).

## 3. Results

Overall, 2,478 participants completed at-least one time point and were included in the study. Cohort 1 included 1,262 (50.9%) participants, and cohort 2 included 1,216 participants (49.1%). 517 participants (20.9%) completed at least two surveys. The mean age of the participants was 34.2 (SD=11.9), and their age ranged between 18 to 91. Of the participants, 1,394 were female (56.2%), 1,474 (59.4%) lived with their spouse and 1,008 (40.7%) lived with at least one of their children. The sample’s sociodemographic properties, stratified by the time of survey’s completion, are presented in Table 1.

**Table 1.** Sociodemographic characteristics of the study sample

| **Variable** | ***T1 (March 20)*** | ***T2 (May 20)*** | ***T3 (Sep 20)*** | ***T4 (Dec 20)*** | ***T5 (June 21)*** |
| --- | --- | --- | --- | --- | --- |
| **N** |  |  |  |  |  |
| *Total* | 1,262 | 241 | 1,460 | 319 | 300 |
| *Cohort 1* | 1,262 | 241 | 244 | 186 | 172 |
| *Cohort 2* | - | - | 1,216 | 133 | 128 |
| **Age** |  |  |  |  |  |
| *Mean (SD)* | 35.3 (12.1) | 37.3 (12.6) | 33.9 (11.7) | 37.1 (12.5) | 36.6 (12.1) |
| *Range* | 18 - 90 | 19 - 78 | 18 - 91 | 19 - 91 | 19 - 75 |
| **Sex** |  |  |  |  |  |
| *Male* | 577 (45.7%) | 75 (31.1%) | 583 (39.9%) | 105 (32.9%) | 107 (35.7%) |
| *Female* | 685 (54.3%) | 166 (68.9%) | 875 (59.9%) | 214 (67.1%) | 193 (64.3%) |
| **Living with spouse** | 764 (60.5%) | 148 (61.4%) | 859 (58.8%) | 198 (62.1%) | 184 (61.3%) |
| **Living with children** | 543 (43.0%) | 107 (44.4%) | 580 (39.7%) | 148 (46.4%) | 140 (46.7%) |
| **Income** |  |  |  |  |  |
| *Below average* | 504 (39.9%) | 111 (46.0%) | 729 (49.9%) | 158 (49.5%) | 139 (46.4%) |
| *Average* | 322 (25.5%) | 59 (24.5%) | 271 (18.6%) | 68 (21.3%) | 64 (21.3%) |
| *Above average* | 408 (32.3%) | 59 (24.5%) | 342 (23.5%) | 76 (23.8%) | 83 (27.7%) |
| **Employment status (before COVID-19)** |  |  |  |  |  |
| *Unemployed* | 81 (6.4%) | 8 (3.3%) | 123 (8.4%) | 13 (4.1%) | 18 (6.0%) |
| *Paid employee* | 716 (56.7%) | 138 (57.5%) | 785 (54.7%) | 182 (57.1%) | 162 (54.4%) |
| *Self-employed* | 109 (8.6%) | 32 (13.3%) | 126 (8.6%) | 37 (11.6%) | 37 (12.4%) |
| *Student* | 242 (19.2%) | 46 (19.2%) | 186 (12.7%) | 50 (15.7%) | 60 (20.1%) |
| *Retired* | 36 (2.9%) | 9 (3.8%) | 26 (1.8%) | 12 (3.8%) | 7 (2.3%) |
| *Military/other* | 78 (6.1%) | 7 (2.9%) | 127 (8.7%) | 14 (4.4%) | 14 (4.7%) |

### 3.1. Trajectories of Depression, Anxiety, PTSS and PRSF

Depression, Anxiety, and PTSS demonstrated significant quadratic trends (χ^*2*^_*(4)*_>57, *p*<.0001) which peaked during the second lockdown, with 50.6% experiencing above-cutoff anxiety, 39.2% experiencing above-cutoff depression, and 28.4% experiencing above-cutoff PTSS (see Figure 2).

**Figure 2.**
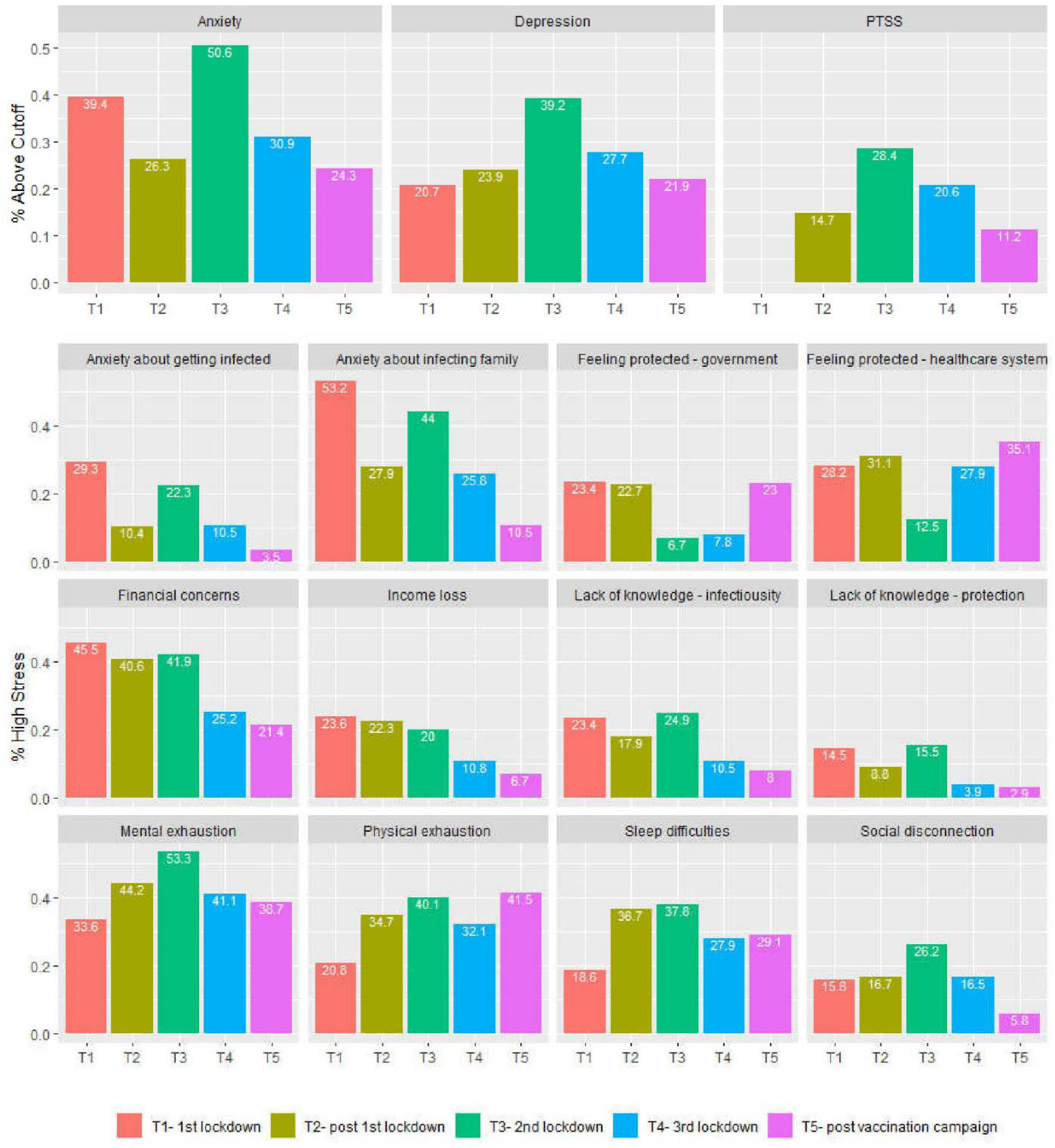
Temporal dynamics of Anxiety, Depression, PTSS and PRSF

The most prevalent levels of high pandemic related stress at T1 were observed for anxiety about infecting family (53.2%), financial concerns (45.5%) and mental exhaustion (33.6%). Financial, health-related and protection PRSFs decreased between T1 and T5, while fatigue measures increased or remained unchanged, including mental exhaustion (from 33.6% to 38.7%), physical exhaustion (from 20.8% to 41.5%) and sleep difficulties (from 18.6% to 29.1%). For all PRSFs, the change in rates over time points was statistically significant in χ^2^ test (*p*<.001). The dynamics of PRSF are depicted in Figure 2.

### 3.2. Time Sensitive Associations of Sociodemographic Factors and PRSF with Anxiety, Depression and PTSS

#### 3.2.1. Sociodemographic factors

Age was the only factor that was negatively associated with both depression (*β*=-0.05, *p*<.001) and anxiety (*β*=-0.07, *p*<.0001), above and beyond specific time points. Female sex associated with higher levels of anxiety (*β*=0.20, *p*<.0001). Depression was negatively associated with several protective factors, including living with children (*β*=-0.12, *p*<.001), living with a spouse (*β*=-0.10, *p*=.004) and having higher income (*β*=-0.05, *p*=.005). PTSS were not associated with any sociodemographic factor (see Table 2).

**Table 2.**
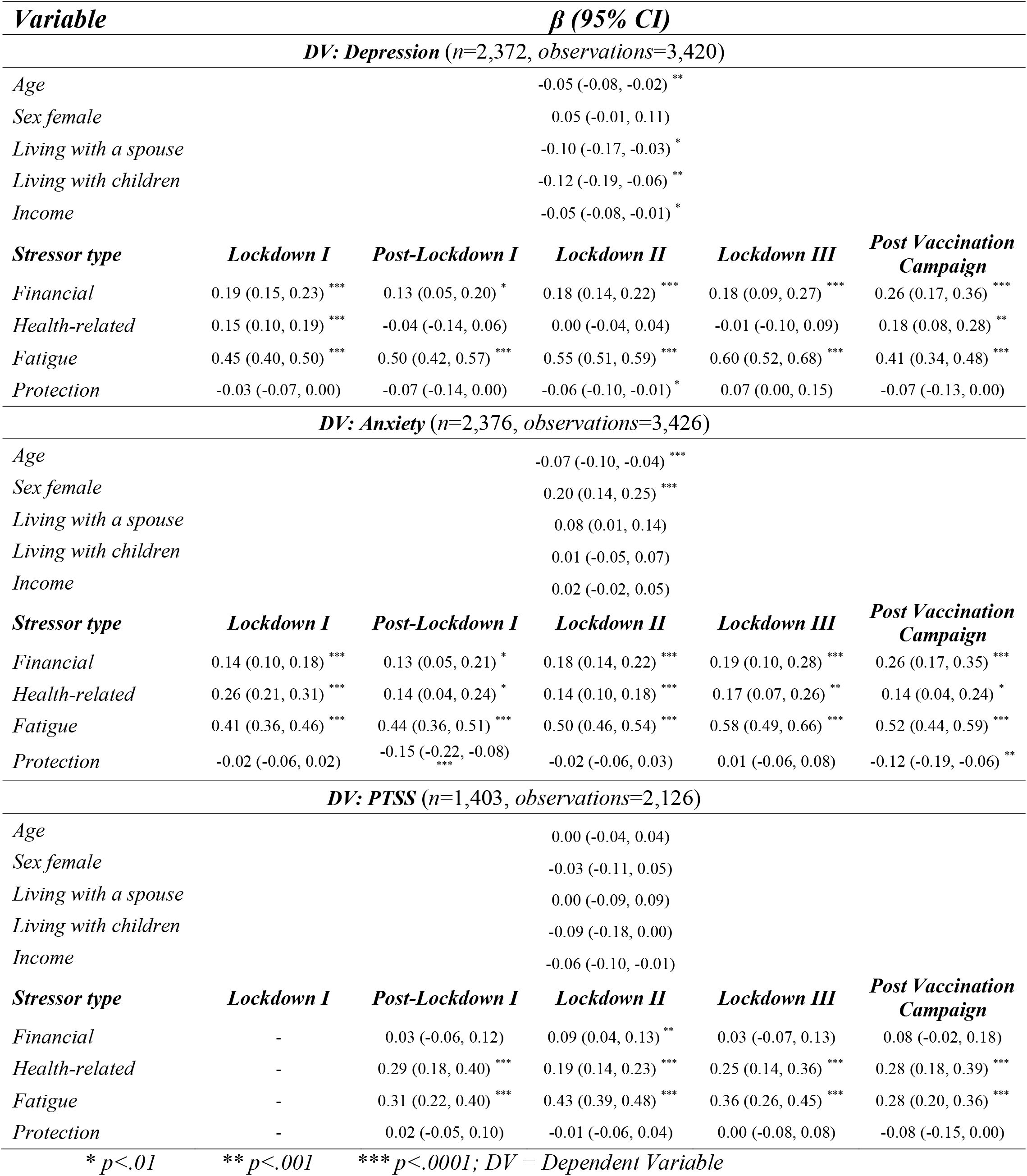
Time-sensitive analysis of cross-sectional data with mixed-effects linear regression

| Variable | | $\beta$ (95% CI) | | | |
| --- | --- | --- | --- | --- | --- |
| DV: Depression (n=2,372, observations=3,420) |  |  |  |  |  |
| Age |  | -0.05 (-0.08, -0.02) ** |  |  |  |
| Sex female |  | 0.05 (-0.01, 0.11) |  |  |  |
| Living with a spouse |  | -0.10 (-0.17, -0.03) * |  |  |  |
| Living with children |  | -0.12 (-0.19, -0.06) ** |  |  |  |
| Income |  | -0.05 (-0.08, -0.01) * |  |  |  |
| Stressor type | Lockdown I | Post-Lockdown I | Lockdown II | Lockdown III | Post Vaccination Campaign |
| Financial | 0.19 (0.15, 0.23) *** | 0.13 (0.05, 0.20) * | 0.18 (0.14, 0.22) *** | 0.18 (0.09, 0.27) *** | 0.26 (0.17, 0.36) *** |
| Health-related | 0.15 (0.10, 0.19) *** | -0.04 (-0.14, 0.06) | 0.00 (-0.04, 0.04) | -0.01 (-0.10, 0.09) | 0.18 (0.08, 0.28) ** |
| Fatigue | 0.45 (0.40, 0.50) *** | 0.50 (0.42, 0.57) *** | 0.55 (0.51, 0.59) *** | 0.60 (0.52, 0.68) *** | 0.41 (0.34, 0.48) *** |
| Protection | -0.03 (-0.07, 0.00) | -0.07 (-0.14, 0.00) | -0.06 (-0.10, -0.01) * | 0.07 (0.00, 0.15) | -0.07 (-0.13, 0.00) |
| DV: Anxiety (n=2,376, observations=3,426) |  |  |  |  |  |
| Age |  | -0.07 (-0.10, -0.04) *** |  |  |  |
| Sex female |  | 0.20 (0.14, 0.25) *** |  |  |  |
| Living with a spouse |  | 0.08 (0.01, 0.14) |  |  |  |
| Living with children |  | 0.01 (-0.05, 0.07) |  |  |  |
| Income |  | 0.02 (-0.02, 0.05) |  |  |  |
| Stressor type | Lockdown I | Post-Lockdown I | Lockdown II | Lockdown III | Post Vaccination Campaign |
| Financial | 0.14 (0.10, 0.18) *** | 0.13 (0.05, 0.21) * | 0.18 (0.14, 0.22) *** | 0.19 (0.10, 0.28) *** | 0.26 (0.17, 0.35) *** |
| Health-related | 0.26 (0.21, 0.31) *** | 0.14 (0.04, 0.24) * | 0.14 (0.10, 0.18) *** | 0.17 (0.07, 0.26) ** | 0.14 (0.04, 0.24) * |
| Fatigue | 0.41 (0.36, 0.46) *** | 0.44 (0.36, 0.51) *** | 0.50 (0.46, 0.54) *** | 0.58 (0.49, 0.66) *** | 0.52 (0.44, 0.59) *** |
| Protection | -0.02 (-0.06, 0.02) | -0.15 (-0.22, -0.08) *** | -0.02 (-0.06, 0.03) | 0.01 (-0.06, 0.08) | -0.12 (-0.19, -0.06) ** |
| DV: PTSS (n=1,403, observations=2,126) |  |  |  |  |  |
| Age |  | 0.00 (-0.04, 0.04) |  |  |  |
| Sex female |  | -0.03 (-0.11, 0.05) |  |  |  |
| Living with a spouse |  | 0.00 (-0.09, 0.09) |  |  |  |
| Living with children |  | -0.09 (-0.18, 0.00) |  |  |  |
| Income |  | -0.06 (-0.10, -0.01) |  |  |  |
| Stressor type | Lockdown I | Post-Lockdown I | Lockdown II | Lockdown III | Post Vaccination Campaign |
| Financial | - | 0.03 (-0.06, 0.12) | 0.09 (0.04, 0.13) ** | 0.03 (-0.07, 0.13) | 0.08 (-0.02, 0.18) |
| Health-related | - | 0.29 (0.18, 0.40) *** | 0.19 (0.14, 0.23) *** | 0.25 (0.14, 0.36) *** | 0.28 (0.18, 0.39) *** |
| Fatigue | - | 0.31 (0.22, 0.40) *** | 0.43 (0.39, 0.48) *** | 0.36 (0.26, 0.45) *** | 0.28 (0.20, 0.36) *** |
| Protection | - | 0.02 (-0.05, 0.10) | -0.01 (-0.06, 0.04) | 0.00 (-0.08, 0.08) | -0.08 (-0.15, 0.00) |
| * $p<.01$ | | ** $p<.001$ | *** $p<.0001$; DV = Dependent Variable | | |

#### 3.2.2. Pandemic-Related Stress Factors

During the first lockdown (T1), both depression and anxiety positively associated with financial stressors (*β* between 0.14 to 0.19, *p*<.0001), health-related stressors (*β* between 0.15 to 0.26, *p*<.0001) and fatigue (*β* between 0.41 to 0.45, *p*<.0001), but not with protection. Financial stressors and fatigue remained significantly associated with depression and anxiety across all time points, with fatigue consistently presenting the strongest association with both. Health-related stressors remained significantly associated with anxiety across all time points (*β* between 0.14 to 0.26, *p*<.01). However, the positive association among health-related stressors and depression vanished after the first lockdown (T2, *β*=-0.04, *p*=.41) and remained insignificant during the second and third lockdowns, only to reappear at the 16-month follow-up (*β*=0.18, *p*<.001). Feelings of protection by authority were negatively associated with anxiety after the first lockdown (T2; *β*=-0.15, *p*<.0001), and at the 16-month follow-up (T5; *β*=-0.12, *p*<.001) (Table 2).

PTSS were first measured after the first lockdown. It associated with fatigue (*β*=0.31, *p*<.0001) and health-related stressors (*β*=0.29, *p*<.001), and remained significant across all time points. Again, fatigue had the strongest association with PTSS across all time points. Financial stressors associated with PTSS only during the second lockdown (*β*=0.09, *p*<.001). (Table 2)

### 3.3. Longitudinal Intercorrelations among changes in PRSF, Depression, Anxiety and PTSS

Worsening in (i.e., increase in) fatigue had the strongest association with worsening of depression (*β*=0.36, *p*<.0001), anxiety (*β*=0.35, *p*<.0001) and PTSS (*β*=0.22, *p*<.0001). Worsening of financial stressors was associated with worsening of depression (*β*=0.18, *p*<.0001) and anxiety (*β*=0.16, *p*<.0001) but not PTSS. Worsening of health-related stressors was associated with worsening in anxiety (*β*=0.11, *p*<.001) and PTSS (*β*=0.16, *p*<.0001) but not depression. Worsening (i.e., decrease in) feelings of protection was associated with worsening in depression (*β*=-0.09, *p*=.003) and anxiety (*β*=-0.16, *p*<.0001), but not PTSS. The results are depicted in Figure 3.

**Figure 3.**
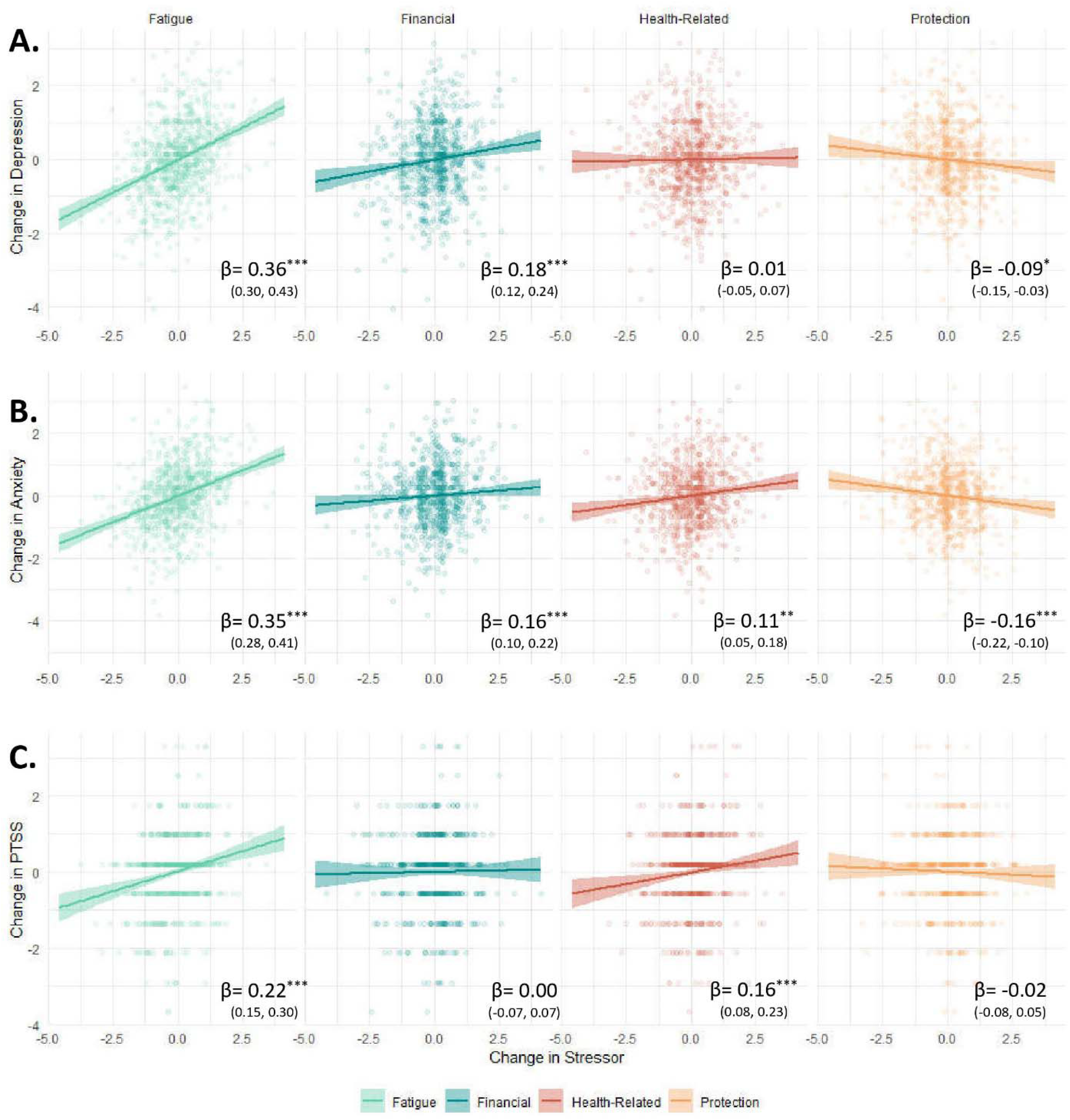
Longitudinal intercorrelations of PRSF with Depression, Anxiety and PTSS *\* p<.01 ** p<.001 *** p<.0001*

## 4. Discussion

In this study we examined the relationship between numerous pandemic-related stressors (financial, health-related, fatigue and sense of protection by the authorities) and psychological distress facets, specifically anxiety, depression and PTSS. Findings showed that financial concerns were associated with depression and anxiety, while health-related concerns were more consistently associated with anxiety and PTSS than with depression. Notably, fatigue was the only stressor that had strong and consistent associations with all three outcomes. We regard these associations as most notable, since they emerged repeatedly across different analyses (observed both cross-sectionally and longitudinally), thus illustrating consistent relations between specific stressors and specific psychopathologies. Sense of protection by authority was only mildly associated with anxiety when examined cross-sectionally, but its deterioration overtime was associated with exacerbation of both anxiety and depression.

As expected, fatigue was the most prominent risk factor for mental health, and the strongest predictor for emotional deterioration in all facets. Put differently, individuals who experienced a depletion in energy and resources as the pandemic progressed were the ones who were most likely to experience an increase in anxiety, depression and PTSS. This robust connection between fatigue and mental health might paint a gloomy picture for some individuals: while most people succeeded in building resilience to health-related concerns, they failed to do the same for fatigue, meaning they could not adapt to the constant feeling of mental and physical exhaustion, sleep problems and sense of social disconnection. This strong association might be partially explained by the centrality of sleep disorders in the pandemic (Mandelkorn et al., 2021). Sleep disruptions have become a central public concern during COVID-19, as they are affected not only by stress and fear but also by the new reality of remote working, layoffs, and lack of outdoor activity, which have disrupted many ‘timekeepers’ that dictated arising and sleeping hours for many people (Morin, Carrier, Bastien, & Godbout, 2020). Together with reduced daylight exposure, it is possible that the pandemic have caused alterations in circadian rhythms for many people, making them un-synchronized with the demands of their day-to-day (Morin et al., 2020), and consequently leading to deterioration of their mental health. Other aspects of fatigue, such as physical exhaustion, can be linked to the global concerns about a potential rise in overweight and obesity during COVID-19 (Clemmensen, Petersen, & Sørensen, 2020). It is possible that lack of physical activity, alterations in eating habits and weight gain have promoted sense of physical exhaustion, which translated to depression and anxiety by both biological mediators (e.g., endocrine abnormalities (Hryhorczuk, Sharma, & Fulton, 2013)) or psychological factors such as decrease in self-esteem (Vittengl, 2018). Another aspect of fatigue, mental exhaustion, can be linked to psychological fatigue and burnout, both concepts whom potential contribution to mental health deterioration was highly discussed during the pandemic (Griffith, 2020; Michie et al., 2020). Our findings emphasize the magnitude of this link, above and beyond many other prominent stressors.

However, the extensive time that passed between each of our surveys (which ranged from several weeks to several months) limit our ability to clearly determine directionality between fatigue and psychiatric morbidity, since the temporal effects of these processes over one another most probably occur much faster. Thus, fatigue might not be solely a stressor, but also a symptom (American Psychiatric Association, 2013) of psychopathology. It is also possible that fatigue was initially triggered by the reality of the pandemic (lockdowns, unemployment, constant worries, etc.), later developing a positive feedback loop with other aspects of the phenomenology of depression, anxiety and PTSS (e.g., sadness, fear), and eventually becoming entrapped in a vicious cycle of exhaustion and mental strain. Future studies can implement time-series analysis on longitudinal data to further clarify the causal association of fatigue with mental health outcomes.

Our study replicated previous findings which tied financial and health related concerns to spiking rates of depression and anxiety at the early stage of the pandemic (Barzilay et al., 2020; Bitan et al., 2020; Hertz-Palmor et al., 2021; Hoffart, Johnson, & Ebrahimi, 2021; Thayer & Gildner, 2021). However, on the span of sixteen months, financial concerns became uniquely correlated with depression, while health-related concerns became prominently associated with PTSS. A possible explanation for this difference might be that financial strain led to degradation of social stance, self-esteem, self-confidence and self-efficacy (as opposed to helplessness), which are known precursors of dysphoria, despair and eventually depression (Blazer, 2002; Lorant et al., 2007; Pryce et al., 2011; Roberts & Monroe, 1994). In contrast, the substantial health threat posed by COVID-19 might have highlighted the possibility of death, arising the potential of health-related fears to translate into traumatic symptomatology (Holbrook, Hoyt, Stein, & Sieber, 2001). These findings are especially interesting since depression and PTSS many times entangle together (O’Donnell, Creamer, & Pattison, 2004).

In accordance with previous reports, our study revealed strong associations of different stressors to mental health symptoms during the first lockdown, (Levy & Cohen-Louck, 2021; Niedzwiedz et al., 2021; Patsali et al., 2020). As time elapsed, we observed a gradual decline of these associations (e.g. the association between health-related concerns and depression), a finding which corresponds with the general recovery of mental health after lockdowns (Fancourt, Steptoe, & Bu, 2021; Picó-Pérez et al., 2021; Prati & Mancini, 2021). Interestingly, several months into the vaccination campaign we observed the re-appearance and strengthening of these associations (e.g., financial concerns with depression and anxiety, health-related concerns with depression). In the case of health-related concerns, the initial strong correlation with depression and anxiety during the first lockdown, followed by their decline during the second and third lockdowns, might be explained by changes in the public opinion on the fatality of the disease, which eased by the second lockdown. Surprisingly, the correlation restored its original strength after the vaccination campaign. This might be due to rumors of the more dangerous delta variant, which were reported to reinstate worries among the public (Alhasan et al., 2021). Notion of this new danger, being even more devastating after the hope of defeating the virus with mass vaccination, might have reinstated the association between fear of contraction and feelings of despair and depression. In contrary, the association of mental health to financial concerns was not influenced by lockdowns, and even increased after the vaccination campaign. This association seemed to be unaffected by fluctuations in the severity of the pandemic situation in the country. This finding validates and elaborates previous findings, showing that the damage that economic hardships place on mental health is consistent and exclusive (Hertz-Palmor et al., 2021).

This study has several limitations. First, the sampling was not random, but rather a “snowball” recruitment. Although by weighting the data we have narrowed the error margins, it does not neutralize the selection bias completely (Pierce, McManus, et al., 2020). Second, although our cross-sectional data relies on a large sample, our longitudinal data relies on a much smaller body due to high rates of attrition. Although this might expose our study to II type errors and biases, to the best of our knowledge our study is still the first endeavor to examine so many participants over many time points in Israel. Considering the relatively small population and the limited body of current studies on COVID-19 in the country, we believe that our data shed much needed light on the situation in Israel during the first year and a half of COVID-19. Lastly, our study employed online crowdsourcing data gathering, using self-report measures with their inherent limitations (Fadnes, Taube, & Tylleskär, 2009). Still, arguably the robustness of the findings we report, replicated for different time points, cohorts and analyses, mitigates most doubts regarding generalizability of our findings.

This study expands our understanding of the unique and dynamic influence of COVID-19 on the public’s mental health. Two years into this ongoing global crisis, we recognize that the mental needs and risks of the public varies at different points in time, and interacts with lockdown orders and the virus’s spread. Our findings emphasize the centrality of fatigue in the conservation and exacerbation of common mental disorders, to a larger extent than other prominent stressors such as financial, health-related concerns, or concerns about protection by the authorities. Most importantly, our findings accentuate the multitude of risk factors for psychiatric morbidity during this complicated epoch, and signals policy makers not to disregard mental health when confronting the virus.

## Supporting information

Supplemental data - weighting procedure

Supplemental data - Exploratory Factor Analysis

## Data Availability

The datasets used and/or analyzed during the current study are available from the corresponding author upon request.

## Acknowledgements

We wish to thank Uzi Dann, Amit Loewenthal and Tanya Hai in their assistance in distributing the surveys. We thank the participants of the study for dedicating their time, again and again, to share their thoughts and feelings during these difficult times.

## Contributors

All authors contributed to, reviewed, and approved the final manuscript. Conceiving and designing the study: *NH-P, NM, MM, SDI, RG, DG, IM-P*

Data collection: *NH-P, NM, DG*

Statistical analysis: *NH-P*

Data interpretation and writing the final manuscript: *NH-P, SR*

Reviewing and editing the final manuscript: *NM, MM, SDI, AA, EM-D, IH-O, RG, DG, IM-P*

## Ethics statement

The study was approved by the Institutional Review Board of Sheba Medical Center, Tel Hashomer, Israel (IRS#SMC-7182-20).

