## Supplemental data - weighting procedure for "A 16-Month Longitudinal Investigation of Risk and Protective Factors for Mental Health Outcomes Throughout Three National Lockdowns and a Mass Vaccination Campaign: Evidence from a Weighted Israeli Sample During COVID-19"

To assure that the data is representative of the Israeli population at the time of the study, the sample was weighted against age and sex distribution in the adult Israeli population in 2020 (18 years or older). The data upon which we based the weighting procedure was taken from the official website of Israel Central Bureau of Statistics (available at <https://www.cbs.gov.il/EN/pages/default.aspx>). Descriptive statistics present the unweighted data characteristics as were observed in the sample, and the regression models present the weighted estimates.

**Table S1**. Age distributions in Israel 2020, separated by sex

| **Overall** | **Sex** | | **Age** |
| --- | --- | --- | --- |
|  | **Male** | **Female** |  |
| **9,289,761** | **4,613,239** | **4,676,522** | **Overall** |
| 176,595 | 90,698 | 85,897 | **0** |
| 182,081 | 93,790 | 88,291 | **1** |
| 185,786 | 95,262 | 90,524 | **2** |
| 185,420 | 95,178 | 90,242 | **3** |
| 183,799 | 94,587 | 89,212 | **4** |
| 181,844 | 93,483 | 88,361 | **5** |
| 179,905 | 92,623 | 87,282 | **6** |
| 175,342 | 90,169 | 85,173 | **7** |
| 174,864 | 89,432 | 85,432 | **8** |
| 170,533 | 87,301 | 83,232 | **9** |
| 170,492 | 87,525 | 82,966 | **10** |
| 166,022 | 84,904 | 81,118 | **11** |
| 160,458 | 81,993 | 78,464 | **12** |
| 156,002 | 79,978 | 76,024 | **13** |
| 152,843 | 78,376 | 74,467 | **14** |
| 148,538 | 76,321 | 72,218 | **15** |
| 148,832 | 76,297 | 72,535 | **16** |
| 149,230 | 76,272 | 72,958 | **17** |
| 144,175 | 73,579 | 70,596 | **18** |
| 141,128 | 72,182 | 68,946 | **19** |
| 141,072 | 72,181 | 68,891 | **20** |
| 136,515 | 69,694 | 66,821 | **21** |
| 135,893 | 69,239 | 66,654 | **22** |
| 131,664 | 67,039 | 64,625 | **23** |
| 129,570 | 65,925 | 63,645 | **24** |
| 125,906 | 64,008 | 61,898 | **25** |
| 126,026 | 63,872 | 62,154 | **26** |
| 125,275 | 63,629 | 61,646 | **27** |
| 124,487 | 62,973 | 61,514 | **28** |
| 123,955 | 62,495 | 61,460 | **29** |
| 122,924 | 61,956 | 60,968 | **30** |
| 122,388 | 61,513 | 60,875 | **31** |
| 121,079 | 60,913 | 60,166 | **32** |
| 119,734 | 59,844 | 59,890 | **33** |
| 120,139 | 60,011 | 60,128 | **34** |
| 119,531 | 59,585 | 59,947 | **35** |
| 119,835 | 59,669 | 60,166 | **36** |
| 120,024 | 60,111 | 59,912 | **37** |
| 117,094 | 58,044 | 59,050 | **38** |
| 114,659 | 56,894 | 57,765 | **39** |
| 114,831 | 56,810 | 58,021 | **40** |
| 113,893 | 56,687 | 57,206 | **41** |
| 110,089 | 54,269 | 55,820 | **42** |
| 112,182 | 55,627 | 56,555 | **43** |
| 115,060 | 57,051 | 58,009 | **44** |
| 112,795 | 55,736 | 57,059 | **45** |
| 108,995 | 54,198 | 54,798 | **46** |
| 105,327 | 51,965 | 53,362 | **47** |
| 102,893 | 50,772 | 52,121 | **48** |
| 102,635 | 50,457 | 52,178 | **49** |
| 98,381 | 48,571 | 49,810 | **50** |
| 93,197 | 45,974 | 47,222 | **51** |
| 89,294 | 44,024 | 45,270 | **52** |
| 83,181 | 40,889 | 42,292 | **53** |
| 84,910 | 41,464 | 43,447 | **54** |
| 83,480 | 41,114 | 42,366 | **55** |
| 81,879 | 40,208 | 41,672 | **56** |
| 79,011 | 38,422 | 40,589 | **57** |
| 78,763 | 37,887 | 40,876 | **58** |
| 77,742 | 37,181 | 40,561 | **59** |
| 80,223 | 38,384 | 41,839 | **60** |
| 77,267 | 36,873 | 40,393 | **61** |
| 75,868 | 36,059 | 39,809 | **62** |
| 74,481 | 35,313 | 39,167 | **63** |
| 73,584 | 34,755 | 38,830 | **64** |
| 74,220 | 34,884 | 39,336 | **65** |
| 71,600 | 33,604 | 37,996 | **66** |
| 71,830 | 33,773 | 38,058 | **67** |
| 71,821 | 33,639 | 38,182 | **68** |
| 70,672 | 32,756 | 37,916 | **69** |
| 69,152 | 32,197 | 36,955 | **70** |
| 61,948 | 28,632 | 33,315 | **71** |
| 60,797 | 27,998 | 32,799 | **72** |
| 65,066 | 29,523 | 35,544 | **73** |
| 58,200 | 26,481 | 31,719 | **74** |
| 45,061 | 20,551 | 24,510 | **75** |
| 37,722 | 17,263 | 20,459 | **76** |
| 32,050 | 14,365 | 17,685 | **77** |
| 29,812 | 13,005 | 16,807 | **78** |
| 30,835 | 13,437 | 17,398 | **79** |
| 30,584 | 12,929 | 17,655 | **80** |
| 29,907 | 12,787 | 17,120 | **81** |
| 29,777 | 12,380 | 17,397 | **82** |
| 28,765 | 11,824 | 16,942 | **83** |
| 24,713 | 10,178 | 14,535 | **84** |
| 21,358 | 8,726 | 12,632 | **85** |
| 18,321 | 7,254 | 11,066 | **86** |
| 15,551 | 6,049 | 9,503 | **87** |
| 14,821 | 5,729 | 9,092 | **88** |
| 11,837 | 4,533 | 7,304 | **89** |
| 11,079 | 4,196 | 6,883 | **90** |
| 8,814 | 3,140 | 5,674 | **91** |
| 7,374 | 2,621 | 4,753 | **92** |
| 5,643 | 1,944 | 3,699 | **93** |
| 4,633 | 1,584 | 3,049 | **94** |
| 3,675 | 1,212 | 2,462 | **95** |
| 2,891 | 912 | 1,979 | **96** |
| 1,977 | 600 | 1,377 | **97** |
| 1,339 | 410 | 929 | **98** |
| 796 | 300 | 496 | **99** |
| 3,498 | 1,588 | 1,910 | **+100** |

**Table S2.** Distribution of the percentage of people on each age group in the actual population and in the sample at each survey point in the cross-sectional analysis

| **Age Group** | **Population** | | **Sample – T1** | | **Sample – T2** | | **Sample – T3** | | **Sample – T4** | | **Sample – T5** | |
| --- | --- | --- | --- | --- | --- | --- | --- | --- | --- | --- | --- | --- |
|  | **Male** | **Female** | **Male** | **Female** | **Male** | **Female** | **Male** | **Female** | **Male** | **Female** | **Male** | **Female** |
| **18-25** | 0.060 | 0.057 | 0.098 | 0.120 | 0.044 | 0.100 | 0.065 | 0.207 | 0.033 | 0.117 | 0.051 | 0.109 |
| **26-35** | 0.066 | 0.066 | 0.193 | 0.199 | 0.124 | 0.259 | 0.150 | 0.210 | 0.126 | 0.264 | 0.137 | 0.256 |
| **36-45** | 0.061 | 0.062 | 0.099 | 0.107 | 0.064 | 0.171 | 0.088 | 0.097 | 0.084 | 0.150 | 0.080 | 0.131 |
| **46-55** | 0.051 | 0.052 | 0.032 | 0.060 | 0.028 | 0.080 | 0.048 | 0.046 | 0.042 | 0.075 | 0.042 | 0.077 |
| **56-65** | 0.040 | 0.043 | 0.015 | 0.030 | 0.020 | 0.028 | 0.017 | 0.011 | 0.015 | 0.009 | 0.019 | 0.019 |
| **65-91** | 0.049 | 0.061 | 0.013 | 0.018 | 0.020 | 0.024 | 0.011 | 0.012 | 0.015 | 0.036 | 0.013 | 0.026 |

**Table S3.** Weights in the cross-sectional analysis (weights above 1 represent under-representation in the sample, weights below 1 represent over-representation)

| **Age Group** | **Sample – T1** | | **Sample – T2** | | **Sample – T3** | | **Sample – T4** | | **Sample – T5** | |
| --- | --- | --- | --- | --- | --- | --- | --- | --- | --- | --- |
|  | **Male** | **Female** | **Male** | **Female** | **Male** | **Female** | **Male** | **Female** | **Male** | **Female** |
| **18-25** | 0.608 | 0.478 | 1.360 | 0.575 | 0.923 | 0.276 | 1.805 | 0.489 | 1.166 | 0.527 |
| **26-35** | 0.344 | 0.329 | 0.538 | 0.253 | 0.443 | 0.312 | 0.526 | 0.248 | 0.483 | 0.256 |
| **36-45** | 0.622 | 0.586 | 0.964 | 0.364 | 0.695 | 0.644 | 0.731 | 0.415 | 0.769 | 0.476 |
| **46-55** | 1.585 | 0.868 | 1.812 | 0.652 | 1.058 | 1.118 | 1.202 | 0.692 | 1.217 | 0.678 |
| **56-65** | 2.696 | 1.468 | 1.999 | 1.556 | 2.373 | 3.953 | 2.652 | 4.816 | 2.078 | 2.263 |
| **65-91** | 3.721 | 3.401 | 2.469 | 2.545 | 4.482 | 5.235 | 3.276 | 1.688 | 3.849 | 2.380 |

**Table S4.** Distribution of the percentage of people on each age group in the actual population and in the sample at each survey point in the longitudinal deltas analysis

| **Age Group** | **Population** | | **Sample – T1-T2** | | **Sample – T2-T3** | | **Sample – T3-T4** | | **Sample – T4-T5** | |
| --- | --- | --- | --- | --- | --- | --- | --- | --- | --- | --- |
|  | **Male** | **Female** | **Male** | **Female** | **Male** | **Female** | **Male** | **Female** | **Male** | **Female** |
| **18-25** | 0.060 | 0.057 | 0.044 | 0.100 | 0.022 | 0.106 | 0.033 | 0.117 | 0.035 | 0.096 |
| **26-35** | 0.066 | 0.066 | 0.124 | 0.260 | 0.100 | 0.306 | 0.127 | 0.265 | 0.106 | 0.278 |
| **36-45** | 0.061 | 0.062 | 0.064 | 0.172 | 0.061 | 0.189 | 0.084 | 0.151 | 0.091 | 0.146 |
| **46-55** | 0.051 | 0.052 | 0.028 | 0.076 | 0.033 | 0.067 | 0.042 | 0.075 | 0.045 | 0.086 |
| **56-65** | 0.040 | 0.043 | 0.020 | 0.028 | 0.022 | 0.017 | 0.015 | 0.012 | 0.015 | 0.010 |
| **65-91** | 0.049 | 0.061 | 0.020 | 0.024 | 0.028 | 0.028 | 0.015 | 0.030 | 0.020 | 0.035 |

**Table S5.** Weights in the longitudinal delta analysis (weights above 1 represent under-representation in the sample, weights below 1 represent over-representation)

| **Age Group** | **Sample – T1-T2** | | **Sample – T2-T3** | | **Sample – T3-T4** | | **Sample – T4-T5** | |
| --- | --- | --- | --- | --- | --- | --- | --- | --- |
|  | **Male** | **Female** | **Male** | **Female** | **Male** | **Female** | **Male** | **Female** |
| **18-25** | 1.355 | 0.573 | 2.683 | 0.543 | 1.799 | 0.488 | 1.686 | 0.597 |
| **26-35** | 0.535 | 0.252 | 0.664 | 0.214 | 0.525 | 0.247 | 0.626 | 0.236 |
| **36-45** | 0.960 | 0.363 | 1.006 | 0.330 | 0.729 | 0.414 | 0.676 | 0.426 |
| **46-55** | 1.805 | 0.684 | 1.516 | 0.780 | 1.198 | 0.690 | 1.112 | 0.605 |
| **56-65** | 1.991 | 1.550 | 1.792 | 2.603 | 2.644 | 3.601 | 2.628 | 4.295 |
| **65-91** | 2.459 | 2.535 | 1.771 | 2.190 | 3.266 | 2.020 | 2.435 | 1.721 |
