## Supplemental data - Exploratory Factor Analysis for "A 16-Month Longitudinal Investigation of Risk and Protective Factors for Mental Health Outcomes Throughout Three National Lockdowns and a Mass Vaccination Campaign: Evidence from a Weighted Israeli Sample During COVID-19"

**Table S6.** Exploratory factor loadings of the Pandemic-Related Stress Inventory (PRSF) + Income loss item

| Cluster | PRSF – In recent days, following the outspread of the coronavirus, I felt… | F1 | F2 | F3 | F4 |
| --- | --- | --- | --- | --- | --- |
| Fatigue | Mental exhaustion | .87 |  |  |  |
|  | Physical exhaustion | .83 |  |  |  |
|  | Sleep difficulties | .79 |  |  |  |
|  | Social disconnection and sense of being shunned by others | .55 |  |  |  |
| Health-related | Lack of knowledge about the virus’ infectiousity and virulence |  | .81 |  |  |
|  | Lack of knowledge about prevention and protection from the virus |  | .78 |  |  |
|  | Anxiety about infecting my family |  | .76 |  |  |
|  | Anxiety about getting infected with the virus |  | .73 |  |  |
| Sense of protection | Protected by the government and local authorities |  |  | 1.00 |  |
|  | Protected by the healthcare system |  |  | 1.00 |  |
| Financial | *[To what extent have you experienced]* Income loss (not an original PRSF item) |  |  |  | .89 |
|  | Financial concerns |  |  |  | .82 |
